# Access to primary care amongst newcomers in Hamilton, Ontario

**DOI:** 10.64898/2025.12.17.25342471

**Authors:** Samuel Ramos-Acevedo, Russell J. de Souza, Nora Abdalla, Sandi M. Azab, Lita Cameron, Mary Crea-Arsenio, Dipika Desai, Deborah D. DiLiberto, Sujane Kandasamy, Patricia Montague, Rosain Stennett, Natalie C. Williams, Gita Wahi, Sonia S. Anand

**Author notes:** **Corresponding Author:** Dr. Sonia Anand, MD, PhD, FRCPC, Associate Vice President, Global Health McMaster University Hamilton, ON, L8S 4K1.

## Abstract

**Background:** More than one in five Canadians (6.5 million people) do not have a family doctor or nurse practitioner they see regularly. Access gaps are greater among immigrants and marginalized populations, who face systemic, cultural, and language barriers to care.

**Objectives:** To evaluate access to primary care provider and dentist among residents of a neighborhood with a high proportion of visible minorities in Hamilton, ON.

**Methods:** Between 2022 and 2024, adults living in Riverdale, a neighbourhood in the city of Hamilton, Ontario in which 51% families were born outside the country, were invited to participate in a cross-sectional survey. Determinants of access to these services were identified using multivariable logistic regression modelling.

**Results:** 930 people completed the survey. Of these, 48% were not born in Canada. The median age of participants was 39 years, with a median time living in Canada of 28 years. Of those who responded, 79.6% had a primary care provider; and 57.1% had a dentist. In multivariable models, living in Canada < 5 years (OR = 0.10; 95% CI: 0.05, 0.20), male sex (OR= 0.56; 95%CI: 0.38, 0.82) and being unmarried (OR = 0.41; 95% CI: 0.27, 0.64) were associated with lower odds of having a primary care provider. Living in Canada for < 5 years (OR = 0.20; 95% CI: 0.11, 0.35), male sex (OR =0.74; 95% CI: 0.55, 0.99), and employment while living below the poverty line (OR=0.50; 95% CI: 0.29, 0.90) were linked to lower access to dental care.

**Conclusion:** In a neighborhood with high proportion of visible minority newcomers in Hamilton, ON, 20% of those surveyed did not have access to a primary care provider, and 43% did not have access to a dentist. Access to primary care was lowest amongst newcomers (within 5 years), men, and those who are unmarried.

## Introduction

A primary health care (PHC) system provides treatment for acute and chronic illnesses, including preventive screening and vaccinations, and is therefore essential for population health (Muldoon et al., 2006). In Canada, the primary care provider, a family doctor or nurse practitioner, is typically the first point of contact and can provide services within scope and refer as needed for specialized care (Kaswa & Von Pressentin, 2025). Most Canadians access care through a primary care provider (PCP) under publicly funded insurance plans (Alemu et al., 2024; Ontario Government., 2025; Ontario. Ministry of health., 2025).

Despite universal healthcare coverage in Canada, gaps persist in PHC. The 2023 Commonwealth Fund survey (Canadian Institute for Health Information, 2023), which found that the percentage of Canadian adults with access to a primary care provider declined from 93% in 2016 to 86% in 2023 (Canadian Institute for Health Information, 2025). However, survey conducted by the Canadian Medical Association in 2025 put the current estimate at 5.9 million, or ∼19.5% of Canadians that do not have a PCP (Canadian Medical Association, 2025), which is an improvement from the Commonwealth surveys, and the CMA’s estimated 6.5 million, or 21.0% in 2022 (Duong & Vogel, 2023), showing the successes of investments and efforts made to fill the gap. People without a PCP seek care more often in emergency departments and ultimately have worse health outcomes than those seen regularly by a PCP (Smithman et al., 2022). New care models in PHC emphasize team-based and community-based approaches, involving physicians, nursing professionals, allied health providers, including psychologists, pharmacists, and community health workers, to improve access and coordination. Alternatively, community health centers and walk-in clinics offer care, but also require a provincial health card (Ministry of Health, 2024; Ontario Ministry of health, 2025).

Moreover, until early 2024, Canada’s publicly-funded PHC system excluded dental care, leaving private insurance, to fill the access gap. This left 32% of Canadians—particularly low-income individuals—without dental coverage (Allison, 2023). Suboptimal dental care increases risk not only for dental problems, but also systemic diseases, including cardiovascular disease (CVD) (Allison, 2023). The new Canadian Dental Care Plan (CDCP) offers federally funded coverage for eligible individuals (Government of Canada, 2024).

Immigration continues to reshape Canada’s demographics, with 395,000 newcomers projected to arrive in 2025–2026 (Government of Canada., 2025). Newcomers, most commonly defined as individuals who have arrived in Canada within the last 5 years, face several barriers to accessing primary care (Pandey et al., 2022; Sundareswaran et al., 2024). Uncertainty regarding eligibility and delays in application or approval processes can hinder timely care at both individual and system levels, resulting in more unmet health needs (Baiden & Evans, 2021; Salami et al., 2019). Permanent residents have provincial coverage, but many temporary residents (e.g. those without a full time job), must rely on private insurance or personal finances to fund care (Ministry of Health, 2024). Refugee claimants, protected persons or people of certain groups have access to temporary healthcare coverage services, through the Interim Federal Health Program (IFHP), until provincial coverage begins (Government of Canada, 2025).

Interrelated individual and systemic barriers such as language barriers, limited knowledge of the health system, and health literacy hinder navigation and communication with providers (Pandey et al., 2022; Sundareswaran et al., 2024). Lower household income, unemployment, long wait times and transportation challenges compound these issues (Pandey et al., 2022; Tsai & Ghahari, 2023).

Moreover, the links between social cohesion, neighbourhood relationships, cultural differences with Canadian health practices and difficulty acculturating lead to discomfort or mistrust in healthcare in newcomer populations, although this is not well captured by current census data (Pandey et al., 2022; Sundareswaran et al., 2024).

The objective of this study was to better understand factors associated with access to primary care and dental services among newcomers. As part of an academic-community-based research program, *Strengthening Community Roots: Anchoring Newcomers in Wellness & Sustainability (SCORE!)*, we conducted a household survey in this neighborhood (Wahi et al., 2023).

## Methods

### Study design and setting

A cross-sectional survey was conducted between 2022 and 2024 in Riverdale, Hamilton, ON—an ethnically-diverse neighbourhood, with 51% of residents born outside Canada, where newcomers (defined as living in Canada for less than 5 years) comprise 16% of the population. The area included two census tracts with ≈ 4,184 dwellings. We divided the tracts by postal code and worked to ensure representative sampling across the areas denoted by the circles shown in **Figure 1**. Ethics approval was obtained from the Hamilton Integrated Research Ethics Board HiREB (15028).

**Figure 1.**
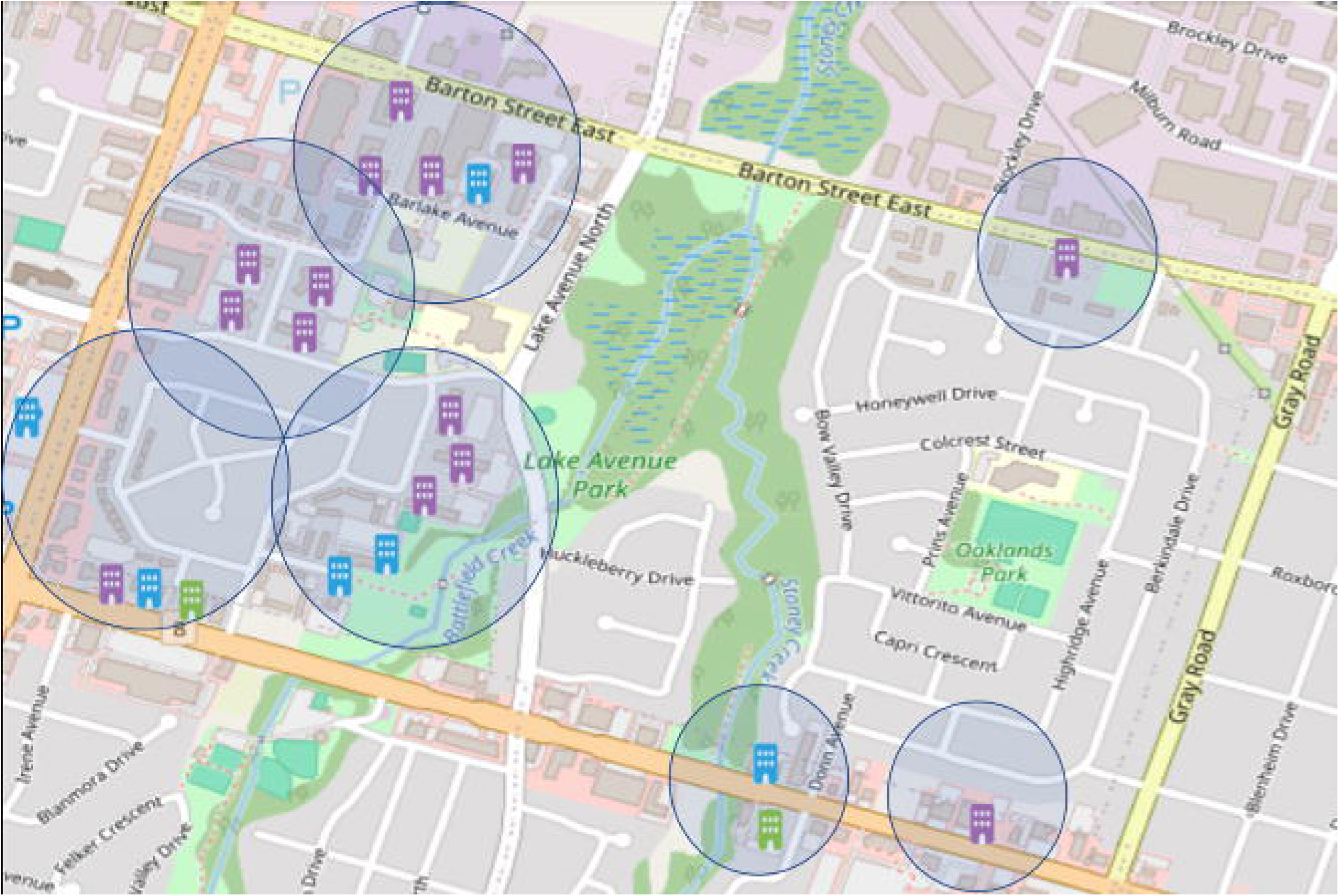
Map representing the geographical area of the Riverdale community and the buildings that were approached, during the study period. (Source: Google My Maps, 2022-2023)

### Recruitment and Eligibility

Postcards with QR codes in five languages were distributed in the neighbourhood. The team also canvassed door-to-door, hosted community events, and promoted the study via social media and local networks. Adults 18 years of age or older living in Riverdale East or West census tracts were eligible, but only one member of a given household could participate in the survey.

### Demographic data

Participants were asked to provide information on age, sex, gender identity, ethnicity, language most often spoken at home, religion, education, marital status, employment, approximated household income after tax, number of years residing in Canada, number of years residing in the Riverdale neighbourhood, and reasons for immigration.

### Household description

Participants answered survey questions that asked to describe their household—type of housing, household ownership, and number of people living in the household.

### Health Literacy

Participants completed the Brief Health Literacy Screen (BHLS) tool, which is a measure of health literacy that has been validated against the Short Test of Functional Health Literacy (S-TOFHLA) (r = 0.42; 95% CI: 0.29, 0.55). The results of the BHLS range from 4 to 20 and were categorized as follows: inadequate (4 – 12), marginal (13 – 16), and adequate (17 – 20) health literacy (Haun et al., n.d.).

### Neighbourhood assessment

Participants answered Likert-style questions designed to assess social cohesion that have been previouslyused in the Neighbourhood Action Strategy in Hamilton, ON (Supplementary Table 1) (City of Hamilton, 2024). Principal component analysis (PCA) was used to reduce a 12-item community Social Determinants of Health (SDoH) (González, 2025) assessment to 4 domains—neighborhood satisfaction (overall perception of and contentment with their residential environment), social cohesion (sense of unity and belonging within a group or society), social capital (resources and benefits individuals gain from their social relationships and networks), and city relationships (residents engagement, and city response to neighbourhood concerns).

### Outcomes

The primary outcome of this analysis was access to a PCP, defined as having a family physician or a nurse practitioner. The secondary outcome was access to a dentist, defined as having a dentist. Both outcome definitions were consistent with Statistics Canada’s definition, with a “yes” answer indicating that the respondent has an existing relationship with 1) a PCP (i.e., family doctor, general practitioner, or nurse practitioner); or 2) a dentist (Canadian Institute for Health Information., 2024).

### Statistical Considerations

#### Sample size

Based on an expected response rate of 25–50% (Harrison et al., 2019), we aimed to collect at least 1,000 surveys from 4,184 private dwellings. At this response rate, we could estimate the proportion of individuals with a primary care provider (PCP) with a precision of ±1.7 to 2.4% and the proportion with a dentist with a precision of ±2.0 to 2.8%, assuming 80% have a PCP and 70% have a dentist.

#### Data Standardization and Summary Measures

Proportions of respondents with access were age- and sex-standardized to Hamilton’s 2021 Census. Continuous variables are summarized as means (SD) or medians (IQR), and categorical variables as frequencies and percentages. Before including in the regression model, some categories were collapsed to save degrees of freedom and improve interpretability: education, marital status, and health literacy, retaining a “Prefer not to answer” option. Time in Canada was grouped as Born in Canada, Long-settled (>10 years), Established newcomer (5–10 years), or Newcomer (<5 years). Poverty status was based on household income and size using the Market Basket Measure for a family of four in Hamilton (Danieles, 2024).

#### Analysis

The SDoH dataset’s suitability for PCA was assessed using Kaiser–Meyer–Olkin test, anti-image matrices, and Cronbach’s α. Factors with eigenvalues ≥1 were retained. Differences in PCP and dentist access were tested using Pearson Χ², and stepwise logistic regression identified predictors, guided by AIC, VIF ≤5, and likelihood-ratio tests. Age, employment, income, marital status, and community SDoH were retained as a priori covariates unless VIF >5. Interaction terms were added to explore subgroup differences, with significant interactions (p<0.05) retained.

## Results

Of 4,184 mailouts, 968 participants initiated the survey and 930 completed it fully (Figure 2). Table 1 shows the sociodemographic characteristics of the population. The respondents’ median age was 39 years (IQR 30-57), and more than half identified as women. Although the survey asked about both sex and gender; most participants preferred to answer the gender question only (“sex” was missing for >95% of respondents; n = 924 missing values). The most common ethnicity was White-European (45.4%), followed by South Asian (21.4%). Over half of respondents were born in Canada, had an education above secondary school, and were employed; nearly half (48.3%) were married or living in common law. The most common type of housing was a privately rented high-rise apartment, home to a median of 3 people (Table 1). Of people who were not born in Canada, (n = 439; 47.7%), the median time lived in Canada was 7 years (IQR 3–20).

**Figure 2.**
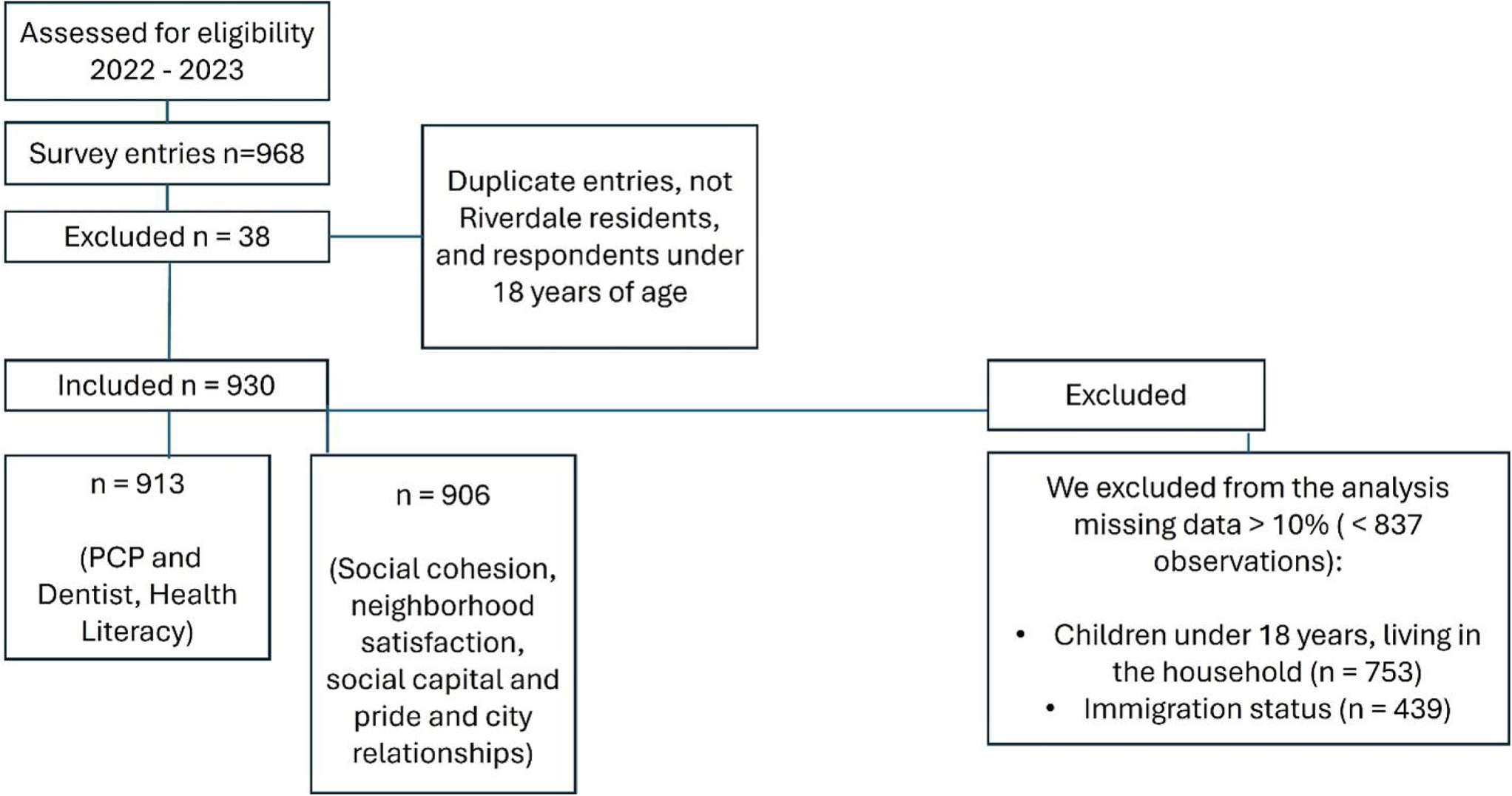
CONSORT Flow diagram of eligible respondents.

**Table 1.** Socio-demographic characteristics of the survey population.

| **Characteristic (n)** | **No. (%) of participants** |
| --- | --- |
| **Gender (n = 921)** |  |
| Man | 393 (42.7) |
| Woman | 515 (55.9) |
| Self-describe | 9 (1.0) |
| Prefer not to answer | 4 (0.4) |
| **Age (n=911)** |  |
| 18-24 | 112 (12.2) |
| 25-34 | 227 (24.7) |
| 35-44 | 208 (22.6) |
| 45-54 | 119 (13.0) |
| 55-64 | 102 (11.1) |
| >65 | 151 (16.4) |
| **Income level (n = 930) #** |  |
| Above MBM, household size | 449 (48.3) |
| Below MBM, household size | 183 (19.7) |
| Prefer not to answer | 298 (32.0) |
| **Employment (n = 921)** |  |
| Employed | 468 (50.8) |
| Unemployed | 236 (25.6) |
| Retired | 173 (18.8) |
| Prefer not to say | 44 (4.8) |
| **Education (n = 921)** |  |
| Secondary school or lower | 390 (42.4) |
| Above secondary school | 513 (55.7) |
| Prefer not to answer | 18 (2.0) |
| **Marital status (n = 921)** |  |
| Married or common law | 445 (48.3) |
| Divorced, separated, or widowed | 431 (46.8) |
| Prefer not to answer | 45 (4.9) |
| **Time in Canada category (n = 921)** |  |
| **Born in Canada** | 482 (52.3) |
| Long settled newcomer (> 10 years in Canada) | 183 (19.9) |
| **Established newcomer (5 – 10 years)** | 101 (11.0) |
| **Newcomer (< 5 years)** | 155 (16.8) |
| **Ethnicity (n = 921)** |  |
| White European | 422 (45.4) |
| Indigenous | 18 (1.9) |
| South Asian | 199 (21.4) |
| Black | 71 (7.6) |
| Central, South, and Latin America | 17 (1.8) |
| Southeast Asian | 25 (2.7) |
| West and Middle East Asian | 82 (8.8) |
| East Asian | 15 (1.6) |
| Other | 47 (5.1) |
| Prefer not to answer | 43 (4.6) |
| **Language spoken at home (n = 930)** |  |
| English | 671 (72.2) |
| Other than English | 259 (27.9) |
| **Type of housing (n = 930)** |  |
| House (single detached, semi-detached, duplex, townhouse) | 180 (19.4) |
| Low-rise apartment (less than 5 stories) | 70 (7.5) |
| High-rise apartment (more than 5 stories) | 675 (72.7) |
| Mobile home, hotel, rooming house, or group home | 3 (0.3) |
| Other | 1 (0.1) |
| **Rent or Own home (n = 930)** |  |
| Own | 125 (13.4) |
| Rent privately | 730 (78.5) |
| Rent from government | 35 (3.8) |
| Other | 32 (3.4) |
| Prefer not to answer | 8 (0.9) |
| **Religion (n = 921)** |  |
| None | 224 (24.3) |
| Atheist/Agnostic | 31 (3.4) |
| Buddhism | 16 (1.7) |
| Christianity | 313 (34.0) |
| Hinduism | 63 (6.8) |
| Islam | 179 (19.4) |
| Judaism | 1 (0.1) |
| Sikhism | 0 |
| Traditional Chinese | 48 (5.2) |
| Other | 46 (5.0) |
| **Have a PCP (n=913)** |  |
| Yes | 727 (79.6) |
| **Have a Dentist (n=913)** |  |
| Yes | 521 (57.1) |
| **Health literacy (n=913)** |  |
| Adequate and Marginal | 769 (84.2) |
| Inadequate | 144 (15.8) |
| **Community SDoH (n = 906 responding)** |  |
| *I am satisfied with my neighbourhood as a whole* | 819 (90.4) |
| *Living in this neighbourhood gives me a sense of pride* | 791 (88.0) |
| *People in my neighbourhood can be trusted* | 792 (87.4) |
| *People in my neighbourhood share the same values* | 769 (84.9) |
| *My neighbourhood continually looks for solutions to problems rather than being satisfied with the way things are* | 759 (83.8) |
| *I have influence over what my neighbourhood is like* | 769 (84.9) |
| *Residents are invited to be involved in decision making in my neighbourhood* | 698 (77.0) |
| *The city is responsive to residents' inquiries, input, and/or requests* | 740 (81.7) |
| # = Poverty line, based on Provisional Market Basket Measure (MBM) thresholds for the reference family (2 adults, 2 children) (<54,000.00 CAD), and equivalences for other household sizes (square root equivalence scale) for 2024. Source: Statistics Canada (Danieles, 2024).<br>PCP= Primary Care Provider (Family physician or nurse practitioner).<br>SDoH: Social Determinants of Health. |  |

Health literacy was adequate across the population, with a BHLS scale median of 18 (IQR 15–20) and there was strong internal reliability (Cronbach’s α = 0.82). Internal agreement within community SDoH domains was high: neighborhood satisfaction (0.83), social cohesion (0.82), social capital (0.71), and city relationships (0.77).

Of the 930 respondents, 913 completed the question for the primary outcome, of which 727 (79.6%) reported having access to a PCP. Men had lower access to PCP (73.5%) than women (84.2%), and those with a PCP were older than those without (42 years vs. 33 years, p = <0.01) (Table 2). After age- and sex-standardization, men still had lower access to a PCP (78.1% vs. 84.1%). Of those without a PCP (n = 186), 72.0% belonged to a visible minority group, defined as someone who is not an Indigenous person, and who is non-Caucasian in race or non-white in colour (Government of Canada, 2016). A total of 69.3% were born outside of Canada; and 65.6% spoke a language other than English at home (Table 2). Moreover, 31.7% were unemployed, and 18.8% had low health literacy (Supplementary Table 1).

**Table 2.** Sociodemographic and economic characteristics of the surveyed population who have access to a primary care provider.

|  | Have PCP<br>n=913 |  |  |
| --- | --- | --- | --- |
| Characteristic (n) | No. (%) of participants |  |  |
|  | Yes = 727<br>(79.6) | No = 186<br>(20.4) | P-value |
| **Gender (n = 913)** |  |  |  |
| Men | 286 (38.3) | 103 (55.4) | <0.01 |
| Woman | 432 (59.4) | 81 (43.6) | <0.01 |
| Self-describe | 6 (0.8) | 2 (1.1) | 0.38 |
| Prefer not to answer | 3 (0.4) | 0 (0.0) | 0.74 |
| **Age (n = 911)** |  |  |  |
| 18-24 | 77 (10.6) | 35 (18.3) | 0.01 |
| 25-34 | 158 (21.8) | 64 (35.0) | <0.01 |
| 35-44 | 156 (21.5) | 51 (27.5) | 0.08 |
| 45-54 | 105 (14.5) | 12 (6.5) | <0.01 |
| 55-64 | 95 (13.1) | 7 (3.8) | <0.01 |
| >65 | 135 (18.6) | 16 (8.7) | <0.01 |
| **Income level <sup>#</sup> (n = 930)** |  |  |  |
| Above MBM, household size | 370 (50.9) | 70 (37.6) | <0.01 |
| Below MBM, household size | 150 (20.6) | 31 (16.7) | 0.23 |
| Prefer not to answer | 207 (28.5) | 85 (45.7) | <0.01 |
| **Employment (n = 913)** |  |  |  |
| Employed | 366 (52.2) | 97 (52.2) | 0.66 |
| Unemployed | 176 (24.2) | 59 (31.7) | 0.03 |
| Retired | 154 (21.2) | 19 (10.2) | 0.01 |
| Prefer not to say | 31 (4.3) | 11 (5.9) | 0.34 |
| **Education (n = 913)** |  |  |  |
| Secondary school or lower | 311 (42.8) | 76 (40.9) | 0.64 |
| Above secondary school | 403 (55.4) | 107 (57.5) | 0.60 |
| Prefer not to answer | 13 (1.8) | 3 (1.6) | 0.87 |
| **Marital status (n = 913)** |  |  |  |
| Married or common law | 333 (45.8) | 95 (51.1) | 0.19 |
| Divorced, separated, or widowed | 360 (49.5) | 82 (44.1) | 0.20 |
| Prefer not to answer | 34 (4.7) | 9 (4.8) | 0.93 |
| **Time in Canada (n = 913)** |  |  |  |
| Born in Canada | 421 (57.9) | 57 (30.7) | <0.01 |
| Long settled newcomer (> 10 years in Canada) | 153 (21.1) | 28 (15.1) | 0.07 |
| Established newcomer (5 – 10 years) | 75 (10.3) | 26 (14.0) | 0.18 |
| Newcomer (< 5 years) | 78 (10.7) | 75 (40.3) | <0.01 |
| **Ethnicity (n = 913)** |  |  |  |
| White European | 367 (50.5) | 52 (28.0) | <0.01 |
| Indigenous | 15 (2.1) | 3 (1.6) | 0.69 |
| South Asian | 135 (18.6) | 61 (32.8) | <0.01 |
| Black | 51 (7.0) | 20 (10.8) | 0.09 |
| Central, South, and Latin America | 14 (1.9) | 3 (1.6) | 0.78 |
| Southeast Asian | 20 (2.8) | 5 (2.7) | 0.96 |
| West and Middle East Asian | 57 (7.8) | 25 (13.4) | 0.02 |
| East Asian | 10 (1.4) | 5 (2.7) | 0.20 |
| Other | 33 (4.5) | 8 (4.3) | 0.83 |
| Prefer not to answer | 38 (5.2) | 9 (4.8) | 0.89 |
| **Language spoken at home (n = 913)** |  |  |  |
| English | 185 (25.5) | 64 (34.4) | 0.01 |
| Other than English | 542 (74.6) | 122 (65.6) | 0.01 |
| **Type of housing (n = 913)** |  |  |  |
| House (single detached, semi-detached, duplex, townhouse) | 146 (20.1) | 28 (15.1) | 0.11 |
| Low-rise apartment (less than 5 stories) | 61 (8.4) | 8 (4.3) | 0.06 |
| High-rise apartment (more than 5 stories) | 518 (71.3) | 148 (79.6) | 0.02 |
| Mobile home, hotel, rooming house, or group home | 1 (0.1) | 2 (1.1) | 0.04 |
| Other | 1 (0.1) | 0 (0.0) | 0.61 |
| **Home ownership (n = 913)** |  |  |  |
| Own home | 107 (14.7) | 16 (8.6) | 0.02 |
| Rent privately | 562 (77.3) | 155 (88.3) | 0.07 |
| Rent from government | 24 (3.3) | 10 (5.4) | 0.18 |
| Other | 28 (3.9) | 3 (1.6) | 0.13 |
| Prefer not to answer | 6 (0.8) | 2 (1.1) | 0.74 |
| **Religion (n = 913)** |  |  |  |
| None | 183 (25.2) | 38 (20.4) | 0.18 |
| Atheist/Agnostic | 27 (3.7) | 4 (2.2) | 0.29 |
| Buddhism | 9 (1.2) | 7 (3.8) | 0.02 |
| Christianity | 264 (36.3) | 47 (25.3) | 0.01 |
| Hinduism | 40 (5.5) | 22 (11.8) | 0.01 |
| Islam | 131 (18.0) | 47 (25.3) | 0.03 |
| Judaism | 0 (0.0) | 1 (0.5) | 0.05 |
| Sikhism | 30 (4.1) | 17 (9.1) | 0.01 |
| Traditional Chinese | 0 (0.0) | 0 (0.0) | - |
| Other | 43 (5.9) | 3 (1.6) | 0.02 |

PCP= Primary Care Provider (Family physician or nurse practitioner)

Over 70% of participants reported having a high sense of neighbourhood satisfaction, social cohesion, social capital-pride, and good relationships with municipal officials. Social cohesion was similar amongst people born outside of Canada and amongst people born in Canada (4.47 ± 1.13, versus 4.28 ± 1.37 = p = 0.12). In contrast, social capital was lower among people born in Canada than among people born outside Canada (2.17 ± 1.15 vs 2.36 ± 1.08; p<0.01) (Supplementary Table 2).

In multivariable models (Table 3), residing in Canada for less than five years (OR = 0.10; 95% CI: 0.05–0.20), being a man (OR = 0.56; 95% CI: 0.39–0.82), and being single (OR = 0.41; 95% CI: 0.27–0.63) predicted lower odds of having a primary care provider (PCP). Interactions among employment status, household income, health literacy, and language spoken at home were examined, but none were significant for PCP access; however, individuals with adequate health literacy and who spoke languages other than English at home were more likely to have a PCP. No interactions between education and health literacy, nor with newcomer status, income, or employment were identified. Higher social capital was linked to lower odds of having a PCP (OR = 0.80; 95% CI: 0.64–0.99), while other community-level social determinants showed no associations.

**Table 3.** Multivariable analysis of having a primary care provider in the Riverdale community, Hamilton, ON (n=902).

| **Regression variable** |  |
| --- | --- |
| **Gender** | **OR (95%CI)** |
| Woman | Ref |
| Men | 0.56 (0.38, 0.82) |
| Prefer not to answer | 0.92 (0.13, 6.40) |
| Prefer to self-describe | - |
| **Employment and Income per household for a family reference, Hamilton, ON. #** | **OR (95%CI)** |
| Employed, above the poverty line | Ref |
| Employed, below the poverty line | 1.02 (0.50, 2.10) |
| Employed, preferred not to answer about poverty line | 0.90 (0.50, 1.63) |
| Unemployed, above the poverty line | 1.55 (0.66, 3.65) |
| Unemployed, below the poverty line | 1.84 (0.80, 4.25) |
| Unemployed, preferred not to answer about poverty line | 0.58 (0.32, 1.06) |
| Retired, above the poverty line | 1.80 (0.77, 4.22) |
| Retired, below the poverty line | 1.15 (0.29, 4.80) |
| Retired, preferred not to answer about poverty line | 1.33 (0.49, 3.62) |
| Preferred not to answer about employment, above the poverty line | 1.09 (0.25, 4.80) |
| Preferred not to answer about employment, below the poverty line | 1.58 (0.30, 8.34) |
| Preferred not to answer about employment, nor about poverty line | 0.43 (0.12, 1.53) |
| **Education** | **OR (95%CI)** |
| Above secondary school | Ref |
| Lower than secondary school | 0.89 (0.59, 1.34) |
| Prefer not to answer | 0.63 (0.14, 2.69) |
| **Marital status** | **OR (95%CI)** |
| Married/Common law | Ref |
| Single/Divorce/Separated/Widowed | 0.41 (0.27, 0.64) |
| Prefer not to answer | 0.50 (0.19, 1.31) |
| **Time in Canada** | **OR (95%CI)** |
| Born in Canada | Ref |
| Long settled newcomer (> 10 years in Canada) | 0.58 (0.32, 1.05) |
| Established newcomer (5 – 10 years) | 0.29 (0.14, 0.58) |
| Newcomer (< 5 years) | 0.10 (0.05, 0.20) |
| **Ethnicity** | **OR (95%CI)** |
| White ethnicity | Ref |
| Other than white | 0.94 (0.54, 1.64) |
| **Health literacy (BHLS) and the language spoken at home** | **OR (95%CI)** |
| Adequate, speaking English at home | Ref |
| Adequate, speaking other language at home | 2.09 (1.23, 3.54) |
| Inadequate, speaking English at home | 1.28 (0.62, 2.65) |
| Inadequate, speaking other language at home | 1.70 (0.77, 3.75) |
| **Type of house** | **OR (95%CI)** |
| House (single detached, semi-detached, duplex, townhouse) | Ref |
| Low-rise apartment (less than 5 stories) | 1.72 (0.67, 4.50) |
| High-rise apartment (more than 5 stories) | 0.79 (0.48, 1.33) |
| Mobile home, hotel, rooming house, or group home | 0.12 (0.01, 2.31) |
| **Religion** | **OR (95%CI)** |
| Christianity | Ref |
| Other than Christianity | 0.72 (0.47, 1.09) |
| **Community Social Determinants of Health** | **OR (95%CI)** |
| Social cohesion (factor 1) | 1.08 (0.91, 1.29) |
| Social capital (factor 2) | 0.80 (0.64, 0.99) |
| OR: Odds Ratio; CI: Confidence Interval; Ref: Reference group |  |
| Adjusted by: All variables adjusted for the others listed; i.e., Time in Canada, self-identified gender, ethnicity, education, marital status, employment, type of house, religion, poverty line (MBM) (for a family reference in Hamilton, ON), health literacy, language spoken at home, social cohesion (factor 1 (Q1 – Q5), social capital (factor 2 = Q6 – Q9). |  |
| # Poverty line, based on Hamilton Market Basket Measure (MBM) thresholds for the reference family (2 adults, 2 children) (<54,000.00 CAD), and estimated equivalences for other household sizes (square root equivalence scale). Source: Statistics Canada (Danieles, 2024). |  |
| BHLS: Brief Health Literacy Screen |  |

For dental care, 57.1% of respondents reported having a dentist (Supplementary Table 3), with lower prevalence among men compared to women (53.3% vs. 65.0%). Lack of a dentist was more common among those born outside Canada and living in the country for 5–10 years (OR = 0.35; 95% CI: 0.20–0.62), newcomers within 5 years (OR = 0.20; 95% CI: 0.11–0.35), men (OR = 0.74; 95% CI: 0.55–0.99), and individuals employed but living below the poverty line (OR = 0.50; 95% CI: 0.29–0.90) (Table 4). Interactions between health literacy and language spoken at home were not statistically significant, and neither social cohesion nor social capital were associated with dental access (Supplementary Table 4).

**Table 4.** Multivariable analysis of having a Dentist in the Riverdale community in Hamilton, ON (n = 905).

| **Regression variable** | **OR (95% CI)** |
| --- | --- |
| **Gender** |  |
| Woman | Ref |
| Men | 0.73 (0.55, 0.99) |
| Prefer not to answer | 0.26 (0.05, 1.50) |
| Prefer to self-describe | 0.41 (0.03, 5.00) |
| **Employment and Income per household for a family reference, Hamilton, ON. #** |  |
| Being employed with income above the poverty line | Ref |
| Employed with income below the poverty line | 0.50 (0.29, 0.90) |
| Employed, prefer not to answer | 0.99 (0.59, 1.64) |
| Unemployed with income above the poverty line | 0.81 (0.44, 1.52) |
| Unemployed with income below the poverty line | 0.77 (0.43, 1.39) |
| Unemployed, prefer not to answer | 0.58 (0.34, 1.00) |
| Retired with income above the poverty line | 0.57 (0.32, 0.96) |
| Retired with income below the poverty line | 0.28 (0.10, 0.75) |
| Retired, prefer not to answer | 0.45 (0.25, 0.82) |
| Preferred not to answer about employment; above the poverty line | 0.26 (0.07, 0.95) |
| Preferred not to answer about employment; below the poverty line | 0.23 (0.06, 0.82) |
| Preferred not to answer about employment nor about poverty line | 2.17 (0.63, 7.58) |
| **Education** |  |
| Above secondary school | Ref |
| Lower than secondary school | 0.77 (0.56, 1.07) |
| Prefer not to answer | 0.40 (0.12, 1.35) |
| **Marital status** |  |
| Married/Common law | Ref |
| Single/Divorce/Separated/Widowed | 1.01 (0.74, 1.39) |
| Prefer not to answer | 1.25 (0.58, 2.74) |
| **Time in Canada** |  |
| Born in Canada | Ref |
| Long settled (> 10 years in Canada) | 0.77 (0.50, 1.18) |
| Established newcomer (5 – 10 years) | 0.35 (0.20, 0.62) |
| Newcomer (< 5 years) | 0.20 (0.11, 0.35) |
| **Ethnicity** |  |
| White ethnicity | Ref |
| Other than white | 0.67 (0.44, 1.01) |
| **Health literacy (BHLS) and language spoken at home** |  |
| Adequate, speaking English at home | Ref |
| Adequate, speaking other language at home | 0.97 (0.50, 1.89) |
| Inadequate, speaking other language at home | 0.84 (0.43, 1.64) |
| Inadequate, speaking English at home | 0.53 (0.24, 1.19) |
| **Type of house** |  |
| House (single detached, semi-detached, duplex, townhouse) | Ref |
| Low-rise apartment (less than 5 stories) | 0.84 (0.44, 1.59) |
| High-rise apartment (more than 5 stories) | 0.84 (0.60, 1.25) |
| Mobile home, hotel, rooming house, or group home | 2.12 (0.17, 27.12) |
| Other | - |
| **Religion** |  |
| Christianity | Ref |
| Other than Christianity | 1.16 (0.84, 1.60) |
| **Community Social Determinants of Health** |  |
| Social Cohesion (Factor 1) | 1.00 (0.88, 1.15) |
| Social Capital (Factor 2) | 0.91 (0.78, 1.06) |
| OR: Odds Ratio; CI: Confidence Interval; Ref: Reference group.<br>Adjusted by: Time in Canada, self-identified gender, ethnicity, education, marital status, type of house, religion, employment, poverty line (MBM) (for a family reference in Hamilton, ON), health literacy, language spoken at home, social cohesion (factor 1 (Q1 – Q5), social capital (factor 2 = Q6 – Q9).<br># Poverty line, based on Hamilton Market Basket Measure (MBM) thresholds for the reference family (2 adults, 2 children) (<54,000.00 CAD), and estimated equivalences for other household sizes (square root equivalence scale). Source: Statistics Canada (Danieles, 2024).<br>BHLS: Brief Health Literacy Screen. |  |

## Discussion

In this survey of access to PCP and dental care in a neighbourhood with a high proportion of visible minority newcomers, 20% of those surveyed did not have a PCP, and 43% did not have a dentist. Certain groups, such as men, unmarried individuals, and newcomers (within 5 years), were less likely to have a PCP. Those living below the poverty line were less likely to have a dentist. Access to both PCP and a dentist increased with longer time in Canada. These data reinforce the call for greater PCP or other models of primary care access along with reinforcing the urgency of publicly funded dental care (Canadian Institute for Health Information, 2025).

Although this study showed high PCP access (84%), Canada’s rate has declined from 93% in 2016 to 86% in 2023, ranking last among 10 high-income countries in the Commonwealth Fund survey (Canadian Institute for Health Information., 2023). Young single men and newcomers (within 5 years in the country) were notably underserved. Similar patterns occur in Germany and the UK (Carruthers et al., 2023; Tillmann et al., 2018). In Germany, immigrants, first- and second-generation men, younger adults, urban residents, and those with low German proficiency had lower PCP access (Tillmann et al., 2018). In the UK, primary care is free, yet the inability to prove immigration status or pay upfront costs remains a barrier (Asif & Kienzler, 2022). For example, to improve care for immigrants living in the UK during COVID-19, community co-designed programs offered testing and vaccination to everyone without requiring provision of an NHS number, showing that such barriers can be overcome in times of crisis (Burns et al., 2022).

Social and demographic factors compound inequitable access to PCP and dental care in Canada. Women report higher access to PCPs than men (87% vs. 79%) (Canadian Medical Association, 2025). Traditional masculine norms often emphasize self-reliance, which can discourage men from engaging in health-seeking behaviors, such as visiting a healthcare provider for preventive care, or timely treatment. This avoidance increases the likelihood of unrecognized and unmanaged health risks (Bonell et al., 2023). Highly educated immigrants face systemic labour market barriers that push them into low-wage, precarious jobs, and limit access to stable healthcare, factors which may erode social ties (Lam & Triandafyllidou, 2024). Credential devaluation and employment insecurity undermine well-being, particularly among older immigrant men who migrated as adults (Islam & Gilmour, 2023; Lam & Triandafyllidou, 2024). This group reports higher levels of loneliness than age-matched Canadian-born peers. These intersecting vulnerabilities call for socially integrative healthcare approaches that go beyond clinical access to foster connection and trust.

Income and employment status further influence access to care. Some unemployed individuals living below the poverty line access PCPs through community programs or the Interim Federal Health Program (IFHP); however, dental care remains neglected due to competing needs such as food, housing, and essential medical expenses, which results in dental care being perceived as a privilege (Sano & Antabe, 2022). The Canadian Dental Care Plan (CDCP), introduced in early 2025 for residents with net incomes under $90,000, aims to address this gap. Nevertheless, precarious employment—especially among part-time workers on temporary permits—often excludes individuals from provincial coverage like OHIP, leaving many reliant on private insurance or out-of-pocket payments. Nearly one-third of working Canadians lack supplemental insurance, limiting access to uncovered services such as prescriptions (Statistics Canada, 2024). Employer-sponsored drug coverage is most common, yet newcomers (within 5 years) are less likely to benefit compared to Canadian-born workers (Statistics Canada, 2024).

Previous studies link a greater sense of belonging to better healthcare access (Vassilev et al., 2011), and strong social connections to effective resource use, particularly for managing chronic conditions and implementing community-based interventions (Hendryx et al., 2002; Vassilev et al., 2011). Although most participants reported high neighborhood satisfaction, social cohesion, and pride, these community-level determinants did not predict access to care. This challenges assumptions that strong networks improve service uptake and underscores the need for system-level solutions beyond fostering social capital. Social capital scores were higher among individuals born outside Canada, reflecting strong bonding social capital—close ties with family, friends, or similar identities. In multivariate analysis, higher social capital was linked to lower odds of having a PCP, suggesting reliance on community resources rather than formal healthcare. This highlights the need to bridge social capital between and across diverse groups, including those born in the host country.

Health literacy and language are interconnected predictors of primary care access. Effective communication requires sufficient language proficiency to understand treatment options and make informed decisions (Nutbeam & Lloyd, 2021; Schillinger, 2021). Newcomers who spoke a language other than English at home but had adequate health literacy were more likely to have a PCP than those with inadequate literacy. Inadequate health literacy often intersects with social disparities, including lower employment and education levels, and is more prevalent among visible minorities.

Although health literacy was adequate in our sample, associations with care access were limited, except where adequate literacy coexisted with speaking a non-English language at home. This suggests navigation depends on more than language fluency or health literacy alone and may be facilitated by cultural or community brokers. System navigators and community health workers help bridge these gaps, improving outcomes and reducing pressure on PCPs (Pinto et al., 2021).

New care models often face resistance and sustainability challenges within bureaucracies, yet adapting to diverse contexts is essential in countries like Canada. Cultural competence remains critical, but many providers report inadequate training in culturally appropriate care (Turin et al., 2021). Immigrants frequently seek providers who share their cultural and linguistic background, highlighting the need for tailored care (Tsai & Ghahari, 2023). Access to professional interpretation and language support is fragmented, though translating health information and using virtual interventions in multiple languages have improved outcomes and adherence (Mahdavi et al., 2025). Oral health and primary care training should integrate cultural awareness and benefit from streamlined credential recognition, while expanding programs for Canadian PHC students can mitigate shortages. Collaborative frameworks such as community-based participatory research (CBPR) foster trust and co-design culturally relevant solutions; when embedded in health system planning, CBPR enhances service uptake and builds long-term resilience. Looking ahead, Canada’s primary care reforms must integrate these community-engaged approaches with accountability mechanism to ensure that immigrant and low-income populations benefit equitably from system-wide transformation.

## Strengths and Limitations

This study’s strengths include its large sample size, standardized measures, and translation into five commonly spoken neighborhood languages, with multilingual recruiters engaged (Wahi et al., 2023). Limitations include its cross-sectional design, which precludes causal inference, and potential social desirability bias. Although 90% of the target sample was achieved, limited ethnic stratification and missing immigrant category data (53%) restricted subgroup analysis. Access to care facilities and non-physician providers were not assessed. Despite these constraints, the study offers valuable insights into immigrant health access. Time since arrival emerged as the strongest predictor, emphasizing the need for timely, targeted interventions for newcomers. Findings also contribute to understanding social cohesion, social capital, and health literacy among marginalized populations, providing a lens to evaluate Canada’s progress toward equitable care in high-immigration communities such as Riverdale.

## Conclusions

In a neighborhood with a high proportion of visible minority newcomers, 20% of survey respondents did not have access to a primary care provider (PCP) and 43% did not have a dentist, which is lower than the country-wide averages. Newcomers (within 5 years), males, and those who were single experienced lower access to primary care. These findings underscore the urgent need for health system reform for newcomers that prioritizes prevention of chronic illness, strengthens access to primary and dental care with the goal of reducing health disparities in Canada.

## Acknowledgments

The SCORE! team gratefully acknowledges the contributions and efforts of the community members, families, community partners and local organization leaders in Riverdale whose efforts were integral to the success of this research. This work was funded by the Public Health Agency of Canada in the form of a grant to SSA [2223-HQ-000007], and by the Juravinski Research Institute and McMaster Children’s Hospital & McMaster University Department of Pediatrics through support awarded to GW. SSA received funding for a post-doctoral fellow from Novartis and GW received graduate student funding from the Faculty of Health Sciences (McMaster University). SSA is supported by the Heart and Stroke Foundation Chair in Population Health. The funders had no role in study design, data collection and analysis, decision to publish, or preparation of the manuscript.

## Data Availability Statement

Requests for data may be made to the corresponding author. As this is one project of several within the overall funded work, data may not be available until all funded studies are completed.

## Roles

**Samuel Ramos-Acevedo-** methodology, writing original draft, writing – review and editing

**Russell J. de Souza-** funding acquisition, investigation, methodology, writing – review and editing

**Nora Abdalla-** data collection, writing – review and editing

**Sandi Azab- writing –** review & editing

**Lita Cameron- writing –** review & editing

**Mary Crea Arsenio -** funding acquisition, investigation, methodology, writing – review and editing

**Dipika Desai -** writing – review and editing

**Deborah D. DiLiberto -** funding acquisition, investigation, methodology, writing – review and editing

**Sujane Kandasamy:** Conceptualization, Funding acquisition, Investigation, Methodology, Writing – original draft, Writing – review & editing

**Patricia Montague:** Project administration, Writing – review & editing

**Rosain Stennett:** Project administration, Writing – review & editing

**Natalie Williams:** Data management, Project administration, Writing – review & editing

**Gita Wahi:** Conceptualization, Funding acquisition, Investigation, Methodology, Writing – review & editing

**Sonia S Anand:** Conceptualization, Funding acquisition, Investigation, Methodology, Supervision, Writing – review & editing

## SCORE! Research Team

### Investigators

Sonia S. Anand, Shrikant I. Bangdiwala, Andrea Baumann, Jeffrey Brook, Mary Crea-Arsenio, Russell J. de Souza, Dipika Desai, Deborah Diliberto, Kathy Georgiades, Fatimah Jackson-Best, Sujane Kandasamy, Matthew Kwan, K. Bruce Newbold, Diana Sherifali, Amanda Sim, Gita Wahi.

### Operations team

Patty Montague (Program Manager), Natalie Williams, Madison Fach, Abdul Naebi.

### In-Kind Supporters

The Canadian Urban Environmental Health Research Consortium (CANUE); The Chanchlani Research Centre, McMaster University; The Global Health Program, McMaster University; Green Venture; Master of Public Health Program, McMaster University; McMaster Okanagan Committee; Neighbourhood Development, Healthy and Safe Communities Department, City of Hamilton; School of Health and Community Services, Mohawk College; The Starfish Canada; Today’s Family; Infant and Child Health (INCH) Lab, Brock University; the McMaster Evidence Review and Synthesis Team (MERST); Research Institute of St. Joe’s Hamilton; Trees for Hamilton; The Ron Joyce Children’s Health Centre; Facility Services, McMaster University; and the YMCA of Hamilton, Burlington & Brantford.

### Community Partners

Community Action Program for Children (CAPC) (c/o Ghanwa Afach); Environment Hamilton/Friendly Streets; Hamilton Public Library; Hamilton Wentworth District School Board (c/o Jeff Zwolak, Lake Avenue Elementary School); Hamilton Wentworth Catholic District School Board (c/o Morris Hucal); Immigrant Working Centre (IWC) (c/o Claudio Ruiz, Wasan Mohamad, Rosemary Aswani, Ahlam Mohammed);

### Administration

Loshana Sockalingam, Kathy Stewart.

### Students/trainees

Nora Abdalla, Sandi Azab, Shania Bhopa, Ashfia Chowdhury, Fatima Dawood, Rabbi Fazle, Gurlean Gill, Junaid Habibi, Salima Hemani, Margaret Lo, Baanu Manoharan, Saathana Mathirajan, Pranshu Muppidi, Abeerah Murtaza, Adonis Ng, Natasha Ross, Sohnia Sansanwal, Jaanuni Shanjith, Afraah Shirin, Tyler Soberano, Divya Tamilselvan, Sanya Vij, Aamina Zahid.

### Volunteers

Adan Amer, Albi Angjeli, Krishna Basani, Dania Buttu, Naisha Dharia, Paranshi Gupta, Araash Halani, Sarah Hassan, Senaya Karunarathne, Menhaz Munir, Lennisha Nagalingam, Melissa Pereira, Felicia Liu, Shanelle Racine, Dania Rana, Sachin Sergeant, Rosain Stennett.

## References

Alemu, F. W., Yuan, J., Kadish, S., Son, S., Khan, S. S., Nulla, S. M., Nicholson, K., Wilk, P., Thornton, J. S., & Ali, S. (2024). Social determinants of unmet need for primary care: a systematic review. Systematic Reviews, 13(1), 252. 10.1186/s13643-024-02647-5

Allison, P. J. (2023). Canada’s oral health and dental care inequalities and the Canadian Dental Care Plan. In Canadian Journal of Public Health (Vol. 114, Issue 4, pp. 530–533). Institute for Ionics. 10.17269/s41997-023-00800-6

Asif, Z., & Kienzler, H. (2022). Structural barriers to refugee, asylum seeker and undocumented migrant healthcare access. Perceptions of Doctors of the World caseworkers in the UK. SSM - Mental Health, 2. 10.1016/j.ssmmh.2022.100088

Baiden, D., & Evans, M. (2021). Black African Newcomer Women’s Perception of Postpartum Mental Health Services in Canada. Canadian Journal of Nursing Research, 53(3), 202–210. 10.1177/0844562120934273

Bonell, S., Trail, K., Seidler, Z., Patel, D., Oliffe, J. L., & Rice, S. M. (2023). “There’s No Sewing Classes, There’s No Bedazzling Seminars”: The Impact of Masculinity on Social Connectedness and Mental Health for Men Living in Inner-Regional Australia. Sex Roles, 88(1–2), 52–67. 10.1007/s11199-022-01329-7

Burns, R., Stevenson, K., Miller, A., & Hargreaves, S. (2022). Migrant-inclusive healthcare delivery in the UK: Lessons learned from the COVID-19 Pandemic. National Health Service (NHS) Number, 2, 6. https://doi.org/10.1016/j

Canadian Institute for Health Information. (2023, December 8). International survey shows Canada lags behind peer countries in access to primary health care. Https://Www.Cihi.ca/En/International-Survey-Shows-Canada-Lags-behind-Peer-Countries-in-Access-to-Primary-Health-Care.

Canadian Institute for Health Information. (2024, November 13). Canadians With a Regular Health Provider. . Https://Www150.Statcan.Gc.ca/N1/Pub/82-570-x/2023001/Section3-Eng.Htm.

Canadian Institute for Health Information. (2025, August 6). Better access to primary care key to improving health of Canadians. Https://Www.Cihi.ca/En/Taking-the-Pulse-Measuring-Shared-Priorities-for-Canadian-Health-Care-2024/Better-Access-to-Primary-Care-Key-to-Improving-Health-of-Canadians.

Canadian Institute for Health Information. (2025, September 15). International survey shows Canada lags behind peer countries in access to primary health care. Https://Www.Cihi.ca/En/International-Survey-Shows-Canada-Lags-behind-Peer-Countries-in-Access-to-Primary-Health-Care.

Canadian Medical Association. (2025, December 9). New survey reveals access to primary care growing, but 5.9 million adults in Canada still lack regular doctor. Https://Www.Cma.ca/about-Us/What-We-Do/Press-Room/New-Survey-Reveals-Access-Primary-Care-Growing-59-Million-Adults-Canada-Still-Lack-Regular-Doctor.

Carruthers, E., Dobbin, J., Fagan, L., Humphrey, A., Nagasivam, A., Stevenson, K., Yuan, J.-M., Aldridge, R. W., & Burns, R. (2023). Interventions to improve access to primary care for inclusion health groups in England: a scoping review. The Lancet, 402, S32. 10.1016/S0140-6736(23)02081-0

City of Hamilton. (2024, November 26). Neighborhood development. Https://Www.Hamilton.ca/City-Council/Plans-Strategies/Strategies/Neighbourhood-Development.

Danieles, P. Kevin. (2024). Market basket measure research paper : an analysis of the equivalization method. Statistics Canada = Statistique Canada.

Duong, D., & Vogel, L. (2023). National survey highlights worsening primary care access. *CMAJ*. Canadian Medical Association Journal, 195(16), E592–E593. 10.1503/cmaj.1096049

González, Martínez. (2025). Bioestadística amigable. Elsevier.

Government of Canada. (2024, December 18). Canadian Dental Care Plan. Https://Www.Canada.ca/En/Services/Benefits/Dental/Dental-Care-Plan/Apply.Html.

Government of Canada. (2025, May 5). Interim Federal Health Program: Who is eligible. Https://Www.Canada.ca/En/Immigration-Refugees-Citizenship/Services/Refugees/Help-within-Canada/Health-Care/Interim-Federal-Health-Program/Eligibility.Html.

Government of Canada. (2025, June 30). Notice – Supplementary Information for the 2025-2027 Immigration Levels Plan. Https://Www.Canada.ca/En/Immigration-Refugees-Citizenship/News/Notices/Supplementary-Immigration-Levels-2025-2027.Html.

Harrison, S., Henderson, J., Alderdice, F., & Quigley, M. A. (2019). Methods to increase response rates to a population-based maternity survey: A comparison of two pilot studies. BMC Medical Research Methodology, 19(1). 10.1186/s12874-019-0702-3

Haun, J., Noland-Dodd, V., Varnes, J., Graham-Pole, J., Rienzo, B., & Donaldson, P. (n.d.). Testing the BRIEF Health Literacy Screening Tool.

Hendryx, M. S., Ahern, M. M., Lovrich, N. P., & McCurdy, A. H. (2002). Access to health care and community social capital. Health Services Research, 37(1), 87–103. http://www.ncbi.nlm.nih.gov/pubmed/11949928

Islam, M. K., & Gilmour, H. (2023). Immigrant status and loneliness among older Canadians. Health Reports, 34(7), 3–18. 10.25318/82-003-x202300700001-eng

Kaswa, R., & Von Pressentin, K. (2025). Primary health care strengthening through the lens of healthcare system thinking. South African Family Practice, 67(1). 10.4102/safp.v67i1.6039

Lam, L., & Triandafyllidou, A. (2024). Road to nowhere or to somewhere? Migrant pathways in platform work in Canada. Environment and Planning A, 56(4), 1150–1169. 10.1177/0308518X221090248

Mahdavi, S., Fekri, M., Mohammadi-Sarab, S., Mehmandoost, M., & Zarei, E. (2025). The use of telemedicine in family medicine: a scoping review. In BMC Health Services Research (Vol. 25, Issue 1). BioMed Central Ltd. 10.1186/s12913-025-12449-7

Ministry of Health. (2024, December 17). Apply for OHIP and get a health card. Https://Www.Ontario.ca/Page/Apply-Ohip-and-Get-Health-Card.

Muldoon, L. K., Hogg, W. E., & Levitt, M. (2006). Primary Care (PC) and Primary Health Care (PHC). Canadian Journal of Public Health, 97(5), 409–411. 10.1007/BF03405354

Nutbeam, D., & Lloyd, J. E. (2021). Understanding and Responding to Health Literacy as a Social Determinant of Health. Annu. Rev. Public Health, 42, 2020. 10.1146/annurev-publhealth

Ontario Government. (2025, June 30). Ontario’s Primary Care Action Plan, January 2025. Https://Www.Ontario.ca/Page/Ontarios-Primary-Care-Action-Plan-January-2025.

Ontario Ministry of health. (2025, June 23). Ontario’s Primary Care Action Plan, January 2025. Https://Www.Ontario.ca/Page/Ontarios-Primary-Care-Action-Plan-January-2025.

Pandey, M., Kamrul, R., Michaels, C. R., & McCarron, M. (2022). Identifying Barriers to Healthcare Access for New Immigrants: A Qualitative Study in Regina, Saskatchewan, Canada. Journal of Immigrant and Minority Health, 24(1), 188–198. 10.1007/s10903-021-01262-z

Pinto, R. M., Rahman, R., Zanchetta, M. S., & Galhego-Garcia, W. (2021). Brazil’s Community Health Workers Practicing Narrative Medicine: Patients’ Perspectives. Journal of General Internal Medicine, 36(12), 3743–3751. 10.1007/s11606-021-06730-8

Salami, B., Salma, J., & Hegadoren, K. (2019). Access and utilization of mental health services for immigrants and refugees: Perspectives of immigrant service providers. International Journal of Mental Health Nursing, 28(1), 152–161. 10.1111/inm.12512

Sano, Y., & Antabe, R. (2022). Regular Dental Care Utilization: The Case of Immigrants in Ontario, Canada. Journal of Immigrant and Minority Health, 24(1), 162–169. 10.1007/s10903-021-01265-w

Schillinger, D. (2021). Social Determinants, Health Literacy, and Disparities: Intersections and Controversies. In Health literacy research and practice (Vol. 5, Issue 3, pp. e234–e243). NLM (Medline). 10.3928/24748307-20210712-01

Smithman, M. A., Haggerty, J., Gaboury, I., & Breton, M. (2022). Improved access to and continuity of primary care after attachment to a family physician: longitudinal cohort study on centralized waiting lists for unattached patients in Quebec, Canada. BMC Primary Care, 23(1). 10.1186/s12875-022-01850-4

Statistics Canada. (2024, December 17). Study: Gaps in prescription insurance coverage. Https://Www150.Statcan.Gc.ca/N1/Daily-Quotidien/240110/Dq240110a-Eng.Htm.

Sundareswaran, M., Martignetti, L., & Purkey, E. (2024). Barriers to primary care among immigrants and refugees in Peterborough, Ontario: a qualitative study of provider perspectives. BMC Primary Care, 25(1). 10.1186/s12875-024-02453-x

Tillmann, J., Puth, M. T., Frank, L., Weckbecker, K., Klaschik, M., & Münster, E. (2018). Determinants of having no general practitioner in Germany and the influence of a migration background: Results of the German health interview and examination survey for adults (DEGS1). BMC Health Services Research, 18(1). 10.1186/s12913-018-3571-2

Tsai, P. L., & Ghahari, S. (2023). Immigrants’ Experience of Health Care Access in Canada: A Recent Scoping Review. In Journal of Immigrant and Minority Health (Vol. 25, Issue 3, pp. 712–727). Springer. 10.1007/s10903-023-01461-w

Turin, T. C., Chowdhury, N., Haque, S., Rumana, N., Rahman, N., & Lasker, M. A. A. (2021). Meaningful and deep community engagement efforts for pragmatic research and beyond: Engaging with an immigrant/racialised community on equitable access to care. BMJ Global Health, 6(8). 10.1136/bmjgh-2021-006370

Vassilev, I., Rogers, A., Sanders, C., Kennedy, A., Blickem, C., Protheroe, J., Bower, P., Kirk, S., Chew-Graham, C., & Morris, R. (2011). Social networks, social capital and chronic illness self-management: A realist review. In Chronic Illness (Vol. 7, Issue 1, pp. 60–86). 10.1177/1742395310383338

