## Supplementary material for "Access to primary care amongst newcomers in Hamilton, Ontario"

**Supplementary material to accompany Ramos-Acevedo, et al.**

***Assessing Community Social Determinants of Health***

*Using a Likert-type scale, the* ***Brief Health Literacy Screening Tool (BHLS)*** *(Wallston et al., 2014) asked:*

- *How often do you have someone help you read hospital materials?*
- *How confident are you feeling out medical forms by yourself?*
- *How often do you have problems learning about your medical condition because of difficulty understanding written information?*
- *How often do you have a problem understanding what is told to you about your medical condition?*

Using a Likert-type scale, we measure:

- **Social cohesion**
  (People in my Neighbourhood can be trusted, People in my Neighbourhood share the same values)
- **Neighbourhood satisfaction and pride**
  (I am satisfied with my Neighbourhood as a whole, Living in this Neighbourhood gives me a sense of pride)
- **Social capital**
  (My Neighbourhood continually looks for solutions to problems rather than being satisfied with the way things are, I have influence over what my neighbourhood is like)
- **Neighbourhood–city relations**
  (Residents are invited to be involved in decision making in my Neighbourhood, The city is responsive to residents’ inquiries, input, and/or requests)

**Supplementary Table 1. Health literacy, social cohesion, social capital, neighbourhood satisfaction, city relationships, and having a primary care provider.**

|  | **No. (%) of participants**  **PCP** | **No. (%) of participants**  **PCP** | **OR (95% CI)** |
| --- | --- | --- | --- |
| **Health literacy (BHLS) (n=913)** | **Yes (n=727)** | **No (n=186)** |  |
| Adequate | 618 (85.0) | 151 (81.2) | Ref |
| Inadequate | 109 (14.9) | 35 (18.8) | 0.76 (0.49, 1.15) |
| **Neighbourhood satisfaction (n = 906)** | **Yes = 722 (79.7)** | **No = 184 (20.3)** | **OR (95% CI)** |
| *I am satisfied with my Neighbourhood as a whole* | | | |
| Agree | 649 (89.9) | 73 (10.1) | Ref |
| Disagree | 73 (10.1) | 14 (7.6) | 1.36 (0.75, 2.47) |
| *Living in this Neighbourhood gives me a sense of pride* | | | |
| Agree | 631 (87.4) | 166 (90.2) | Ref |
| Disagree | 91 (12.6) | 18 (9.7) | 1.32 (0.77, 2.26) |
| ***Social Cohesion* (n = 906)** | **Yes = 722 (79.7)** | **No = 184 (20.3)** | **OR (95% CI)** |
| *People in my Neighbourhood can be trusted* | | | |
| Agree | 631 (87.4) | 161 (87.5) | Ref |
| Disagree | 91 (12.6) | 23 (12.5) | 1.00 (0.61, 1.64) |
| *People in my Neighbourhood share the same values* | | | |
| Agree | 613 (84.9) | 156 (84.8) | Ref |
| Disagree | 109 (15.1) | 28 (15.2) | 0.99 (0.63, 1.55) |
| **Social Capital (n = 906)** | **Yes = 722 (79.7)** | **No = 184 (20.3)** | **OR (95% CI)** |
| *My Neighbourhood continually looks for solutions to problems rather than being satisfied with the way things are.* | | | |
| Agree | 158 (83.2) | 158 (85.9) | Ref |
| Disagree | 121 (16.8) | 26 (14.1) | 1.22 (0.77,1.93) |
| *I have influence over what my neighbourhood is like* | | | |
| *Agree* | 515 (71.3) | 145 (78.8) | Ref |
| *Disagree* | 207 (28.7) | 39 (21.2) | 1.49 (1.01, 2.20) |
| **City relationships (n = 906)** | **Yes = 722 (79.7)** | **No = 184 (20.3)** | **OR(95% CI)** |
| *Residents are invited to be involved in decision making in my Neighbourhood* | | | |
| Agree | 546 (75.6) | 152 (82.6) | Ref |
| Disagree | 176 (24.4) | 32 (17.4) | 1.53 (1.00, 2.32) |
| *The city is responsive to residents’ inquiries, input, and/or requests* | | | |
| Agree | 581 (80.5) | 159 (86.4) | Ref |
| Disagree | 141 (19.5) | 25 (13.6) | 1.54 (0.97, 2.44) |
| PCP= Primary Care Provider (Family physician or nurse practitioner).  OR: Odds Ratio; CI: Confidence Interval; Ref: Reference group.  BHLS: Brief Health Literacy Screen, PCP: Primary Care Provider,  Adequate; includes neutral  Χ2 Pearson or Fisher; p<0.05 for difference in mean or distribution between those who have a PCP (“yes”) and those who do not (“no”) | | | |

Supplementary Table 2. Univariate odds ratio for the sociodemographic and economic characteristics of having access to primary care.

| **Predictor** | **OR (95%CI)** |
| --- | --- |
| **Gender** |  |
| Woman | Ref |
| Men | 0.52 (0.37, 0.72) |
| Self-describe | 0.56 (0.11, 2.83) |
| Prefer not to answer | - |
| **Age** |  |
| 18-24 | 0.50 (0.32, 0.77) |
| 25-34 | 0.53 (0.37, 0.75) |
| 35-44 | 0.72 (0.50, 1.04) |
| 45-54 | 2.44 (1.32, 4.55) |
| 55-64 | 3.84 (1.75, 8.43) |
| >65 | 2.42 (1.40, 4.18) |
| **Income level** |  |
| Above MBM, household size | Ref |
| Below MBM, household size | 0.91 (0.57, 1.46) |
| Prefer not to answer | 0.46 (0.32, 0.66) |
| **Employment** |  |
| Employed | Ref |
| Unemployed | 0.79 (0.54, 1.14) |
| Retired | 2.14 (1.26, 3.63) |
| Prefer not to answer | 0.74 (0.36, 1.53) |
| **Education (n = 913)** |  |
| Above secondary school | Ref |
| Lower than secondary school | 1.08 (0.78, 1.50) |
| Prefer not to answer | 1.15 (0.32, 4.11) |
| **Marital Status** |  |
| Married, common law | Ref |
| Single, divorced, separated, widowed | 0.80 (0.57, 1.11) |
| Prefer not to answer | 0.86 (0.40, 1.86) |
| **Time in Canada (Category)** |  |
| Born in Canada | Ref |
| Long settled newcomer (> 10 years in Canada) | 0.73 (0.45, 1.20) |
| Established newcomer (5 – 10 years) | 0.39 (0.23, 0.66) |
| Newcomer (< 5 years) | 0.14 (0.09, 0.21) |
| **Ethnicity** |  |
| White ethnicity | Ref |
| Other than white | 0.38 (0.26, 0.54) |
| **Language spoken at home** |  |
| English | Ref |
| Other than English | 0.65 (0.46, 0.91) |
| **Type of house** |  |
| House | Ref |
| Low-rise apartment | 1.46 (0.63, 3.38) |
| High-rise apartment | 0.67 (0.43, 1.04) |
| Mobile home, hotel, rooming house | 0.09 (0.1, 1.09) |
| Other | - |
| **Home ownership** |  |
| Own home | Ref |
| Rent privately | 0.54 (0.31, 0.94) |
| Rent from government | 0.35 (0.14, 0.88) |
| Other | 1.39 (0.37, 5.12) |
| Prefer not to answer | 0.44 (0.08, 2.41) |
| **Religion (dichotomous)** |  |
| Christianity | Ref |
| Other than Christianity | 0.59 (0.41, 0.85) |
| **Health literacy and speaking a language other than English at home** |  |
| Adequate | Ref |
| Inadequate | 0.76 (0.49, 1.15) |
| **Community SoDH** |  |
| Social Cohesion (Factor 1) | 0.87 (0.73, 1.04) |
| Social Capital (Factor 2) | 0.85 (0.72, 1.00) |
| OR: Odds Ratio; CI: Confidence Interval; Ref: Reference group.  # Poverty line, based on Hamilton Market Basket Measure (MBM) thresholds for the reference family (2 adults, 2 children) (<54,000.00 CAD), and estimated equivalences for other household sizes (square root equivalence scale). Source: Statistics Canada (Danieles, 2024).  SoDH: Social Determinants of Health.  Social Cohesion factor 1= Questions 1 – Questions 5)  Social Capital factor 2 = Questions 6 – Questions 9). | |

Supplementary Table 3. Sociodemographic and economic characteristics of the surveyed population who have access to a Dentist.

|  | Have a Dentist  n=913 | |  |
| --- | --- | --- | --- |
| Characteristic (n) | No. (%) of participants | |  |
|  | Yes = 521 (57.1) | No = 392 (42.9) | P value |
| **Gender (n = 913)** |  |  |  |
| Men | 200 (38.4) | 189(48.2) | <0.01 |
| Woman | 316 (60.6) | 197(50.3) | <0.01 |
| Self-describe | 3 (0.6) | 5(1.3) | 0.26 |
| Prefer not to answer | 2 (0.4) | 1(0.2) | 0.73 |
| **Age (n = 911)** |  |  |  |
| 18-24 | 61 (11.7) | 51 (13.0) | 0.54 |
| 25-34 | 102 (19.6) | 120 (30.7) | <0.01 |
| 35-44 | 118 (22.7) | 89 (22.7) | 0.98 |
| 45-54 | 79 (15.1) | 38 (9.7) | 0.01 |
| 55-64 | 74 (14.2) | 28 (7.1) | <0.01 |
| >65 | 86 (16.5) | 65 (16.6) | 0.98 |
| **Income level ^#^ (n = 930)** |  |  |  |
| Above MBM, household size | 279 (53.6) | 161 (41.1) | <0.01 |
| Below MBM, household size | 86 (16.5) | 95 (24.2) | <0.01 |
| Prefer not to answer | 156 (29.9) | 136 (34.7) | 0.12 |
| **Employment (n = 913)** |  |  |  |
| Employed | 284 (54.5) | 179 (45.7) | <0.01 |
| Unemployed | 119 (22.8) | 116 (29.6) | 0.02 |
| Retired | 100 (19.2) | 73 (18.6) | 0.83 |
| Prefer not to say | 18 (3.5) | 24 (6.1) | 0.06 |
| **Education (n = 913)** |  |  |  |
| Lower than secondary school | 294 (56.4) | 216 (55.1) | 0.91 |
| Above secondary school | 220 (42.2) | 167(42.6) | 0.69 |
| Prefer not to answer | 7 (1.3) | 9 (2.3) | 0.27 |
| **Marital status (n = 913)** |  |  |  |
| Single/Divorced/separated | 262 (50.3) | 166 (42.4) | 0.02 |
| Married/Common law | 234 (44.9) | 208 (53.1) | 0.02 |
| Prefer not to answer | 25 (4.8) | 18 (4.6) | 0.88 |
| **Time in Canada (n = 913)** |  |  |  |
| Born in Canada | 327 (62.8) | 151 (38.5) | <0.01 |
| Long settled newcomer (> 10 years in Canada) | 106 (20.4) | 75 (19.1) | 0.65 |
| Established newcomer (5 – 10 years) | 44 (8.4) | 57 (14.6) | <0.01 |
| Newcomer (< 5 years) | 44 (8.4) | 109 (27.8) | <0.01 |
| **Ethnicity (n = 913)** |  |  |  |
| White European | 204 (48.7) | 131 (31.3) | <0.01 |
| Indigenous | 12 (2.3) | 6 (1.5) | 0.40 |
| South Asian | 83 (15.9) | 113 (28.8) | <0.01 |
| Black | 31 (5.9) | 40 (10.2) | 0.02 |
| Central, South, and Latin America | 10 (1.9) | 7 (1.8) | 0.88 |
| Southeast Asian | 12 (2.3) | 13 (3.3) | 0.35 |
| West and Middle East Asian | 31 (5.9) | 51 (13.0) | <0.01 |
| East Asian | 9 (1.7) | 6 (1.5) | 0.81 |
| Other | 25 (4.8) | 16 (4.1) | 0.06 |
| Prefer not to answer | 33 (6.3) | 14 (3.6) | 0.60 |
| **Language spoken at home (n = 913)** |  |  |  |
| Other than English | 112 (21.5) | 137 (34.9) | <0.01 |
| English | 409 (78.5) | 255 (65.1) | <0.01 |
| **Type of housing (n = 913)** |  |  |  |
| House (single detached, semi-detached, duplex, townhouse) | 107 (20.5) | 67 (17.1) | 0.19 |
| Low-rise apartment (less than 5 stories) | 38 (7.3) | 31 (7.9) | 0.73 |
| High-rise apartment( more than 5 stories) | 374 (71.8) | 292 (74.5) | 0.36 |
| Mobile home, hotel, rooming house or group home | 2 (0.4) | 1 (0.2) | 0.74 |
| Other | 0(0) | 1 (0.2) | 0.25 |
| **Home ownership (n = 913)** |  |  |  |
| Own home | 82 (15.7) | 41 (10.5) | 0.02 |
| Rent privately | 403 (77.3) | 314 (80.1) | 0.31 |
| Rent from government | 12 (2.3) | 22 (5.6) | <0.01 |
| Other | 19 (3.6) | 12 (3.1) | 0.63 |
| Prefer not to answer | 5 (0.96) | 3 (0.8) | 0.76 |
| **Religion (n = 913)** |  |  |  |
| None | 138 (26.5) | 83 (21.2) | 0.06 |
| Atheist/Agnostic | 19 (3.6) | 12 (3.1) | 0.63 |
| Buddhism | 9 (1.7) | 7 (1.8) | 0.95 |
| Christianity | 197 (37.8) | 114 (29.1) | <0.01 |
| Hinduism | 25 (4.8) | 37 (9.4) | <0.01 |
| Islam | 81 (15.6) | 97 (24.7) | <0.01 |
| Judaism | 1 (0.2) | - | 0.39 |
| Sikhism | 14 (2.7) | 33 (8.4) | <0.01 |
| Traditional Chinese | - | - | - |
| Other | 37 (7.1) | 9 (2.3) | <0.01 |
| # = Poverty line, based on Provisional Market Basket Measure (MBM) thresholds for the reference family (2 adults, 2 children) (<54,000.00 CAD), and equivalences for other household sizes (square root equivalence scale). Source: Statistics Canada (Danieles, 2024).  Χ^2^ Pearson p<0.05 for difference in mean or distribution between those who have a Dentist (“yes”) and those who do not (“no”). | | | |

**Supplementary Table 4. Health literacy, social cohesion, social capital, neighbourhood satisfaction, city relationships, and having a Dentist.**

|  | **No. (%) of participants**  **Dentist** | **No. (%) of participants**  **Dentist** |  |
| --- | --- | --- | --- |
| **Health literacy (BHLS) (n=913)** | **Yes = 521 (57.1%)** | **No = 392 (42.9)** | **OR (95% CI)** |
| Adequate-Marginal | 456 (87.5) | 313 (79.8) | Ref |
| Inadequate | 65 (12.5) | 79 (20.2) | 0.56 (0.39, 0.80) |
| **Neighbourhood satisfaction (n=906)** | **Yes = 519 (56.8)** | **No = 387 (42.4)** | **OR (95% CI)** |
| *I am satisfied with my Neighbourhood as a whole n (%).* | | | |
| Agree | 474 (91.3) | 345 (89.1) | Ref |
| Disagree | 45 (8.7) | 42 (10.9) | 0.77 (0.50, 1.21) |
| *Living in this Neighbourhood gives me a sense of pride n (%).* | | | |
| Agree | 448 (86.3) | 349 (90.2) | Ref |
| Disagree | 71 (13.7) | 38 (9.8) | 1.45 (0.95, 2.21) |
| ***Social Cohesion* (n=906)** | **Yes = 519 (56.8)** | **No = 387 (42.4)** | **OR (95% CI)** |
| *People in my Neighbourhood can be trusted n (%).* | | | |
| Agree | 449 (86.5) | 343 (88.6) | Ref |
| Disagree | 70 (13.5) | 44 (11.4) | 1.21 (0.81, 1.81) |
| *People in my Neighbourhood share the same values n (%)* | | | |
| Agree | 430 (82.8) | 339 (87.6) | Ref |
| Disagree | 89 (17.2) | 48 (12.4) | 1.46 (1.00, 2.13) |
| **Social Capital (n=906)** | **Yes = 519 (56.8)** | **No = 387 (42.4)** | **OR (95% CI)** |
| *My Neighbourhood continually looks for solutions to problems rather than being satisfied with the way things are n (%) n (%).* | | | |
| Agree | 432 (83.2) | 327 (84.5) | Ref |
| Disagree | 87 (16.8) | 60 (15.5) | 1.09 (0.76, 1.57) |
| *I have influence over what my neighbourhood is like n (%).* | | | |
| *Agree* | 367 (70.7) | 293 (75.7) | Ref |
| *Disagree* | 152 (29.3) | 94 (24.3) | 1.29 (0.95, 1.74) |
| **City relationships (n=906)** | **Yes = 519 (56.8)** | **No = 387 (42.4)** | **OR (95% CI)** |
| *Residents are invited to be involved in decision making in my Neighbourhood n (%).* | | | |
| Agree | 388 (74.8) | 310 (80.1) | Ref |
| Disagree | 131 (25.2) | 77 (19.9) | 1.35 (0.98, 1.86) |
| *The city is responsive to residents’ inquiries, input, and/or requests n (%).* | | | |
| Agree | 419 (80.7) | 321 (82.9) | Ref |
| Disagree | 100 (29.3) | 66 (17.1) | 1.16 (0.82, 1.63) |
| BHLS: Brief Health Literacy Screen, Adequate; includes neutral,  OR: Odds Ratio; CI: Confidence Interval; Ref: Reference group.  Χ^2^ Pearson p<0.05 for difference in mean or distribution between those who have a PCP (“yes”) and those who do not (“no”). | | | |
